# Neurological manifestations of patients with mild-to-moderate COVID-19 attending a public hospital in Lima, Peru

**DOI:** 10.1101/2021.03.16.21253736

**Authors:** Marco H. Carcamo Garcia, Diego D. Garcia Choza, Brenda J. Salazar Linares, Monica M. Diaz

## Abstract

**Objective:** To determine the prevalence and characteristics of the most common neurological manifestations in Peruvian patients with mild-to-moderate COVID-19.

**Methods:** We conducted a single-center prospective, cross-sectional study at an isolation center functioning as a public acute-care hospital during the COVID-19 pandemic in Lima, the capital city of Peru. This was a convenience sample of patients with acute COVID-19 infection and mild-to-moderate respiratory symptoms who presented for hospital admission between September 25 and November 25, 2020. We interviewed participants and collected demographic, medical history and clinical presentation data; all participants underwent a complete physical and neurological examination. Descriptive statistics and prevalence ratios (PR) with corresponding 95% confidence intervals and p-values were calculated to explore between-groups differences.

**Results:** Of 199 patients with mild-to-moderate COVID-19 enrolled in this study, 83% presented with at least one neurological symptom (mean symptom duration 8 +/-6 days). The most common neurological symptoms were headache (72%), hypogeusia or ageusia (41%), hyposmia or anosmia (40%) and dizziness (34%). Only 2.5% of the cohort had an abnormal neurological examination. The majority (42%) had no prior comorbidities. Presence of at least 1 neurological symptom was independently associated with fever, dyspnea, cough, poor appetite, sore throat, chest tightness or diarrhea, but not with comorbid conditions.

**Conclusions:** This cross-sectional study found that headaches, and smell and taste dysfunction are common among patients presenting with mild-to-moderate acute COVID-19 in Lima, Peru. International longitudinal studies are needed to determine the long-term neurological sequelae of COVID-19 during the acute and post-infectious period.

## Introduction

SARS-CoV-2 virus, the causal agent of the COVID-19 infection, was initially identified in December 2019 and declared the cause of a pandemic by the World Health Organization in March 2020^1^. As of February 2021, more than 90 million cases have been reported with more than 2 million related deaths worldwide^2^. Low and middle income countries (LMIC), particularly Latin America, have felt the repercussions of the pandemic due to factors such as income inequalities and heterogeneous nationwide healthcare access resulting in high mortality rates from COVID-19^3,4^. Peru has reported more than 1 million cases and about 40,000 deaths to-date in a country of 33 million inhabitants^2^. Despite this large infection burden, no research has assessed non-respiratory signs or symptoms of the infection among Peruvians.

Various studies worldwide have reported neurological symptoms attributable to COVID-19, including headaches^5^, dizziness^6^, hyposmia or dysgeusia^7,8^, and more severe sequelae (i.e. stroke^9,10^, Guillain-Barre syndrome^11^, and encephalitis^12^). Varying incidences of these acute neurological syndromes have been reported worldwide^6,13^, with few studies from Latin America^14–16^.

The few publications from Peru to-date are isolated case reports on neurological manifestations of COVID-19^17–19^, but no observational studies have been published. International efforts to collect standardized data across the globe on neurological symptoms and syndromes associated with COVID-19 are underway^20,21^, however, more research is needed to determine the regional burden of neurological illness using concerted reporting efforts. We present the first study of neurological signs and symptoms among hospitalized patients with mild-to-moderate COVID-19 in Peru, and one of the only such studies in Latin America.

## Materials & Methods

### Study Population and Procedures

We conducted a single-center, prospective cross-sectional study at the *Centro de Atención y Aislamiento para COVID-19 Villa Panamericana* (Care and Isolation Center for COVID-19 Patients), an isolation center functioning as an acute-care hospital during the COVID-19 pandemic in Lima, the capital city of Peru. This hospital was created by the *EsSalud* – *Seguro Social de Salud* (EsSalud - Social Health Insurance Institute) of the Peruvian government in response to the challenge of insufficient hospital infrastructure in Lima and to offer more healthcare services to the public during the pandemic. The Villa Panamericana was originally a building complex that was constructed during the Pan American Games of Lima, but was adapted during the pandemic to provide healthcare services and a safe space for voluntary isolation of confirmed or suspected persons with COVID-19 infection (with or without compatible symptoms), as well as their close contacts or household members^22^. Therefore, not all patients had a confirmed diagnosis of COVID-19 in order to be admitted to the hospital.

In our study, we interviewed a convenience sample of patients who were hospitalized between September 25 and November 25, 2020 at the time of their hospital admission. Inclusion criteria were: age greater than 18 years and confirmed COVID-19 infection by real-time reverse transcriptase-polymerase chain reaction (rt-PCR) or by rapid antibody test (either IgM and/or IgG positive). At the time of this evaluation, rt-PCR testing was not widely available in public hospitals in Lima, Peru and antibody testing was considered the diagnostic standard nationwide, thus, our inclusion criteria allowed for positive serological testing^23^. We excluded patients with neurological symptoms attributable to a home medication or a known past or current medical illness (i.e. diabetic neuropathy, migraine), and patients with an oxygen requirement, altered mental status or other symptomatology that interfered with their ability to answer the questionnaire. We excluded patients with severe COVID-19 infection or critical illnesses as defined by the diagnostic and classification criteria issued by the National Institute of Health^24^.

The treating study physician (AVM, BJSL, DDGC, MHCG, RSQF) approached each patient at the bedside on the hospital floors at the time of hospital admission. Following explanation of the study procedures and obtaining verbal consent, a face-to-face interview was performed by the physician. This questionnaire consisted of 30 questions (13 sociodemographic, 17 clinical), addressing sociodemographic characteristics, past medical history and home treatment of conditions, as well as current or past acute clinical symptoms of no more than 14 days duration prior to hospital admission. Neurological symptoms were divided into three categories: symptoms affecting the central nervous system (CNS), peripheral nervous system or musculoskeletal system. Following this interview, a physical examination and neurological examination were performed. Due to limited laboratory testing in the hospital facility and no neuroimaging available, these data were not collected as part of this study.

### Data Collection and Statistical Analyses

Participants were selected using convenience sampling, and approximately 30% of all eligible patients during the study period were enrolled. Data was collected using Magpi^25^, a mobile-based data collection application, and exported to CCV for statistical analysis performed using Epi Info version 7.2.4.0^26^. We presented descriptive statistics, including frequencies and percentages for categorical variables and means and standard deviations (SD) for continuous variables. To explore associations between-groups, we estimated prevalence ratios (PR) and their corresponding 95% confidence intervals (CI) and p-values. Our sample of 165 patients with neurological symptoms and 34 without neurological symptoms had a power of 87% to detect differences of 30% or more of the prevalence of comorbidities or non-neurological symptoms.

### Ethical Considerations

This study was approved by the *Instituto de Evaluación de Tecnologías en Salud e Investigación* (IETSI) COVID-19-specific ethics committee on July 13, 2020. No personal identifiers were collected for this study. All patients provided verbal informed consent prior to enrollment in the study. The authors can make the data set from which this study was derived to any investigator who requests it of the corresponding author.

## Results

### Demographic characteristics of study participants

A total of 205 patients with COVID-19 were enrolled in the study, with 6 patients excluded due to incomplete data; therefore, 199 were included in the analysis. Mean age was 43 (SD 15) years with female patients making up more than half (57%) of the study population. We found that most patients were born in Lima (65%) and the majority of patients lived in the southern districts of Lima (**Table 1**). There were no significant difference between those who had neurological symptoms and those who did not have neurological symptoms in terms of age, sex, birth place or current residence (all p>0.05; data not shown). More than two-thirds of the population included in the analysis (72%) had a positive rapid antibody test and the rest had a positive rt-PCR test; **Table 1**.

**Table 1.** **Demographics of Study Population with acute COVID-19 infection (N=199)**

| **Demographics** | **n (%) or mean (SD)** |
| --- | --- |
| Age | 42.8 (15.1) |
| **Sex** |  |
| Male | 85 (42.7%) |
| Female | 114 (57.3%) |
| **COVID-19 diagnosis** |  |
| Rapid antibody test | 144 (72.4%) |
| Real Time-PCR | 55 (27.7%) |
| **Occupation** |  |
| Housewife | 29 (14.6%) |
| Merchant/retail | 18 (9.1%) |
| Student | 13 (6.5%) |
| Military | 10 (5%) |
| **Birthplace** |  |
| Lima | 128 (64.7%) |
| Outside of Lima | 70 (35.4%) |
| **Residency** |  |
| San Juan de Miraflores | 24 (12.1%) |
| Villa Maria del Triunfo | 20 (10.1%) |
| San Juan de Lurigancho | 20 (10.1%) |
| Los Olivos | 16 (8.1%) |
| Villa el Salvador | 15 (7.6%) |
| Chorrillos | 14 (7.1%) |

Abbreviations: PCR=polymerase chain reaction; SD=standard deviation; SpO2=oxygen saturation.

### Clinical characteristics of the study population

The majority (58%) of the patients in our study had at least one comorbidity. The most common comorbidities were: hypercholesterolemia (12%), followed by hypertension (10%), prior history of tuberculosis or other pulmonary disease (9%) and diabetes (7%). A small proportion of patients had a history of cancer (4%); chronic kidney disease (2%) cerebrovascular disease or stroke (1%). Nearly 10% of the cohort had a history of smoking or were current smokers; **Table 2**. There were no significant differences in frequency of comorbidities between those who presented with neurological symptoms and those who did not (all p>0.05; data not shown).

**Table 2.** **Comorbid conditions of study population with acute COVID-19 infection by self-report (N=199)**

| **Comorbid Conditions**<br>**(self-reported)** | **n (%)** |
| --- | --- |
| No comorbidities | 83 (41.7 %) |
| Hypertension | 20 (10.1%) |
| Treated at home | 14 (70%) |
| Diabetes | 13 (6.5%) |
| Treated at home | 10 (76.9%) |
| Hypercholesterolemia | 24 (12.1%) |
| Coronary heart disease | 3 (1.5%) |
| Current smoker | 15 (7.5%) |
| Cerebrovascular disease or stroke | 2 (1%) |
| Obesity | 3 (1.5%) |
| Prior tuberculosis or other pulmonary disease | 17 (8.5%) |
| Cancer | 8 (4%) |
| Chronic kidney disease | 4 (2%) |
| On dialysis | 4 (100%) |
| Chronic gastritis | 6 (3.0%) |
| HIV | 1 (0.5%) |

### Neurological symptoms of the study population

The majority of patients with COVID-19 infection had at least one neurological symptom (83%), with a mean reported symptom duration of 8 (+/- 6) days since symptom onset. However, only 2.5% of the cohort had an abnormal neurological examination at presentation. We found that the most frequent symptoms were those affecting the CNS, with the most common clinical symptom being headache in more than 70% of the population (**Table 3**). Other common symptoms included: dizziness (34%), nausea or vomiting associated with dizziness (25%) and ataxia (8%). Focal motor (7%) and focal sensory (7%) deficits, impaired consciousness (4%) and seizure (0.5%) were less common. Symptoms affecting the peripheral nervous system were also common with hypogeusia or ageusia (41%) and hyposmia or anosmia (40%) present in many patients. Visual changes (12%) were less common, but of those with visual changes, bilateral visual changes and decreased visual acuity were most common. Neuralgia (10%) affected a smaller proportion of the population with most patients having unilateral leg pain. Myalgias were present in nearly half of the population (46%); **Table 3**.

**Table 3.** **Frequency of neurological symptoms among patients presenting with acute COVID-19 infection (N=199)**

| **Neurological Symptoms and Characteristics** | **n ( % ) or mean (SD)** |
| --- | --- |
| At least 1 neurological symptom | 165 (82.9%) |
| Neurological symptom duration prior to hospital presentation (mean [standard deviation] days) | 8 (6) |
| Abnormal neurological examination at time of hospital admission | 4 (2.5%) |
| **Central Nervous System** |  |
| Headache | 143 (71.9%) |
| Dizziness | 68 (34.2%) |
| Nausea or vomiting associated with dizziness | 49 (24.6%) |
| Impaired consciousness | 7 (3.5%) |
| Ataxia | 15 (7.5%) |
| Seizure* | 1 (0.5%) |
| Focal motor deficit | 14 (7%) |
| Focal sensory deficit | 14 (7%) |
| **Peripheral Nervous System** |  |
| Neuralgia or neuropathy | 19 (9.6%) |
| Face- Unilateral | 0 (0%) |
| Face- Bilateral | 2 (10.5%) |
| Arms- Unilateral | 2 (10.5%) |
| Arms- Bilateral | 1 (5.3%) |
| Legs- Unilateral | 3 (15.8%) |
| Legs- Bilateral | 3 (15.8%) |
| Visual symptoms** | 23 (11.6%) |
| Unilateral | 0 (0%) |
| Bilateral | 15 (65.2%) |
| Deficient color vision | 4 (17.4%) |
| Vision loss | 17 (73.9%) |
| Double vision | 3 (13%) |
| Hypogeusia / ageusia | 81 (40.7%) |
| Hyposmia / anosmia | 80 (40.2%) |
| **Musculoskeletal System** |  |
| Myalgia | 92 (46.2%) |

\*Reported seizure was generalized tonic-clonic
\*\*Patients may have had more than one visual symptom.

### Non-neurological symptoms of the study population

The majority of patients had an adequate oxygenation status, with a mean oxygenation saturation of 97.5% at the time of the study interview (Table 4). Nearly 90% of the population had at least one non-neurological symptom at hospital presentation. About half of the population expressed having the following symptoms: cough (57%), fever (53%), poor appetite (49%), sore throat (48%), chest pain (47%), diarrhea (45%), dyspnea (40%). We found that nearly 10% of the population was asymptomatic, presenting with neither COVID-19-related symptoms nor neurological symptoms (**Table 4**).

**Table 4.**
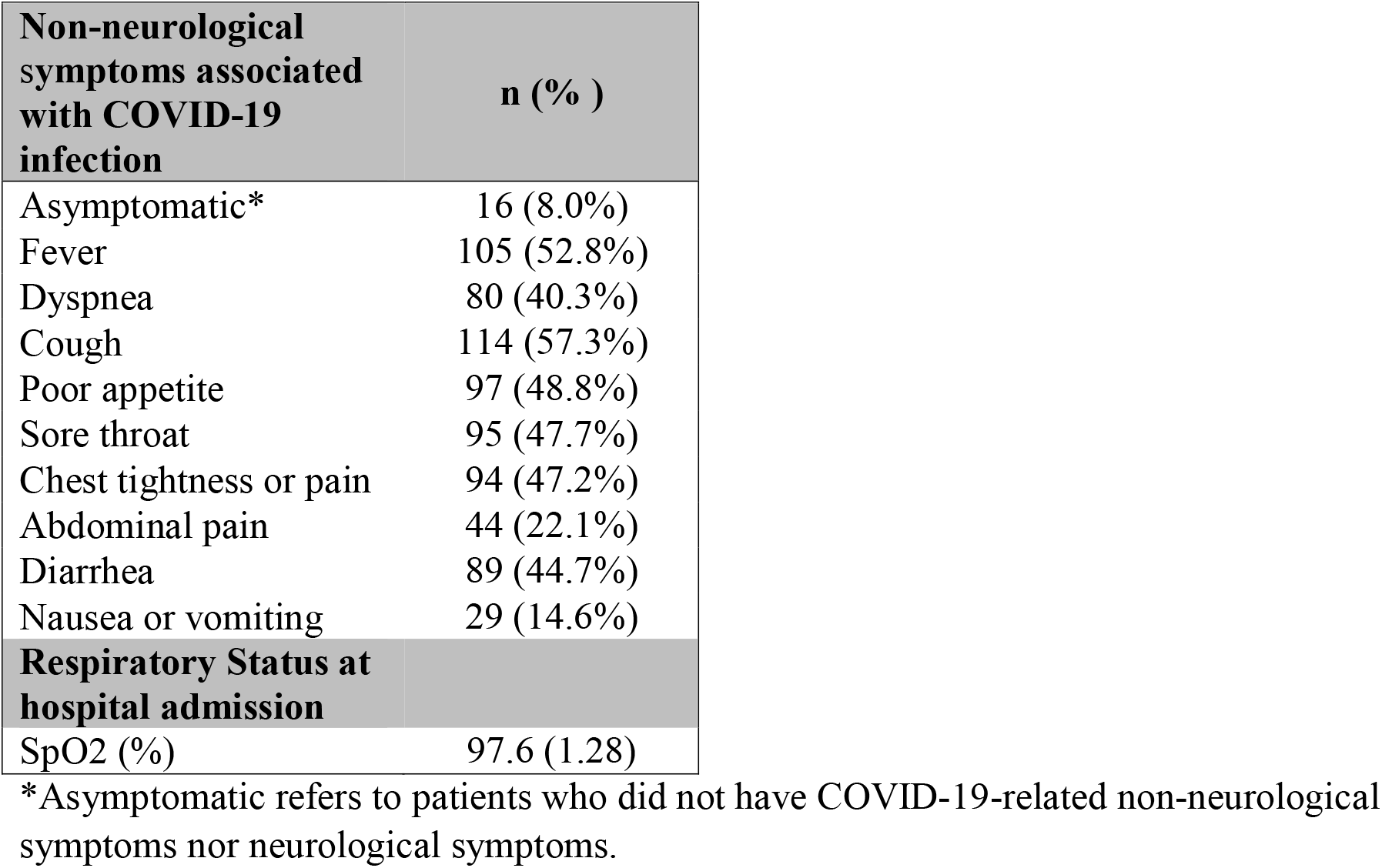
**Frequency of COVID-19-related non-neurological symptoms among the study population (N=199)**

| **Non-neurological symptoms associated with COVID-19 infection** | **n ( % )** |
| --- | --- |
| Asymptomatic* | 16 (8.0%) |
| Fever | 105 (52.8%) |
| Dyspnea | 80 (40.3%) |
| Cough | 114 (57.3%) |
| Poor appetite | 97 (48.8%) |
| Sore throat | 95 (47.7%) |
| Chest tightness or pain | 94 (47.2%) |
| Abdominal pain | 44 (22.1%) |
| Diarrhea | 89 (44.7%) |
| Nausea or vomiting | 29 (14.6%) |
| **Respiratory Status at hospital admission** |  |
| SpO2 (%) | 97.6 (1.28) |

\*Asymptomatic refers to patients who did not have COVID-19-related non-neurological symptoms nor neurological symptoms.

### Relationships between comorbid conditions or non-neurological symptoms and neurological symptoms

Symptoms such as fever, dyspnea, cough, poor appetite, sore throat, chest tightness or pain, diarrhea, and nausea or vomiting were independently associated with having at least one neurological symptom at presentation (all p<0.05). Abdominal pain was not associated with a neurological symptom at hospital admission (**Table 5**). Having a history of a comorbid condition was not associated with having at least one neurological symptom (all p>0.05) (**Table 6)**.

**Table 5.** **Relationship of COVID-19-related symptoms with presence of neurological symptoms among persons with acute COVID-19 infection**.

| **COVID-19-related symptoms** | **PR (95% CI)** | **p value\*** |
| --- | --- | --- |
| Fever | 1.31 (1.14 -1.50) | 0.000 |
| Dyspnea | 1.27 (1.13 -1.43) | 0.000 |
| Cough | 1.24 (1.07 -1.43) | 0.002 |
| Poor appetite | 1.29 (1.14 -1.47) | 0.000 |
| Sore throat | 1.22 (1.08 -1.39) | 0.002 |
| Chest tightness or pain | 1.34 (1.18 -1.52) | 0.000 |
| Abdominal pain | 1.13 (1.00 -1.27) | 0.171 |
| Diarrhea | 1.25 (1.11 -1.41) | 0.001 |
| Nausea or vomiting | 1.20 (1.08 -1.33) | 0.033 |

\*Fischer's exact test
Abbreviations: CI= confidence interval; PR=prevalence ratio

**Table 6.**
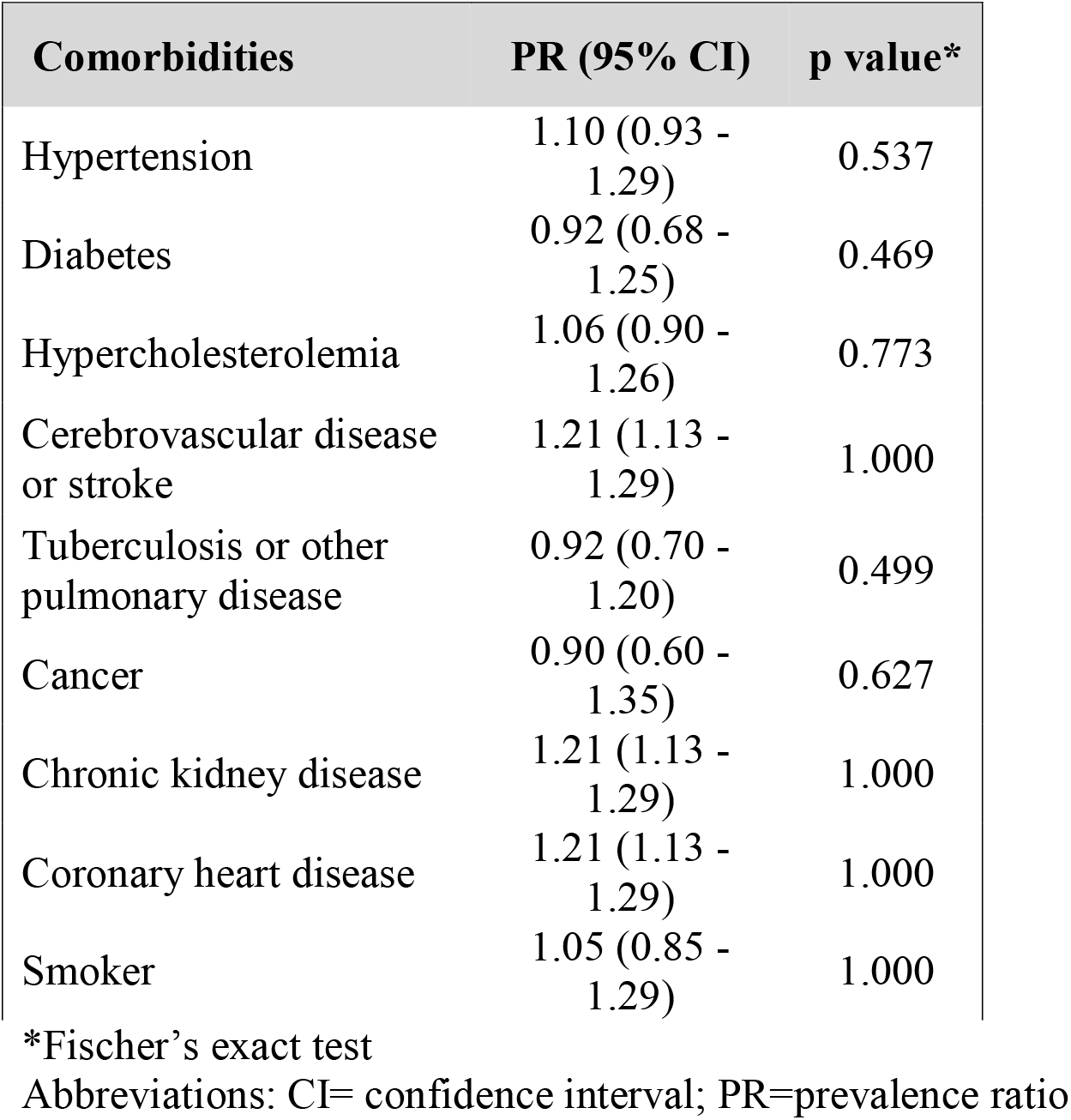
**Relationship of comorbid conditions with presence of neurological symptoms among persons with acute COVID-19 infection**

## Discussion

Our study has demonstrated that patients living in Lima, Peru with acute COVID-19 infection and mild-to-moderate respiratory symptoms have a high frequency of having at least one neurological symptom, with headache, hyposmia/anosmia or hypogeusia/ageusia being the most common. We found that having a prior history of cardiovascular or lung disease was not associated with increased risk of presenting with a neurological symptom, and that having a non-neurological symptom, such as fever, dyspnea, or cough was associated with neurological symptom presentation. We present the first study in Peru to have documented the incidence of neurological symptoms in the setting of mild-to-moderate COVID-19 and one of the few in Latin America. The prevalence of these neurological sequelae associated with COVID-19 has been demonstrated in other countries, but it is important to make a concerted effort to determine their burden worldwide.

Neurological symptom prevalence in the setting of COVID-19 infection thus far has varied widely. In one meta-analysis of 409 patients with SARS-CoV-2 infection and neurological symptoms, neurological alterations varied from 17% to 36%, and the most common neurological symptoms were headache in 17%, dizziness (14%), and altered consciousness (11%)^27^. In our study, we found that more than 80% of patients presenting to this public hospital in the Peruvian capital city of Lima with either a positive COVID-19 rapid antibody or rt-PCR test had at least one neurological symptom with headache being the most common, followed by hyposmia, hypogeusia and dizziness. More debilitating neurological symptoms, such as focal motor or sensory deficit or seizures were rare in our population. Therefore, we have demonstrated an elevated prevalence of these symptoms, which may be attributable to an increased inflammatory response to the infection or sociocultural differences in symptom reporting compared with other countries.

Headache has been widely reported in patients with COVID-19 infection, particularly among those with mild symptoms. Some studies have reported a lower prevalence of headache compared with our study and have not found a relationship between COVID severity and presence of headache. For example, in a meta-analysis of 86 studies (n=14,275), the pooled prevalence of headache was much lower than that of our study (10.1%), with no significant difference in headache prevalence among severe/critically-ill vs. non-severe COVID-19 patents (p=0.78)^28^. Similar to the population in our study, one survey study from Turkey of patients with COVID-19 infection treated in an outpatient setting, found that new-onset headaches occurred in 33% of the population, and more common among those with COVID-19 compared to controls^29^. Our study has demonstrated a higher prevalence of headache than both of these studies, demonstrating the potential utility of headache as a clinical symptom of COVID-19 infection.

Other studies have demonstrated a similar prevalence of headache to that of our study, such as a study from Brazil reporting a headache prevalence of 64%, and about 40% of headaches presenting with anosmia^16^. Contrary to the findings of these authors, we did not find a relationship between the presence of headache and hyposmia in our study. However, in another study from France of mostly mild COVID-19 cases, the headache prevalence was nearly 60% and most headaches had resolved months after infection^5^. Thus, headache is a prevalent feature of COVID-19 infection in many countries and may be a robust indicator of presence of COVID-19 infection among those in whom the diagnosis is questionable or who are awaiting testing results.

The pathophysiology of headache in the setting of COVID-19 has been explored. In some headache syndromes unrelated to COVID-19, cytokine and chemokine release may trigger nociceptive sensory neurons^30^. A similar pain mechanism exists related to COVID-19 infection in which massive release of systemic cytokines and chemokines by macrophages leads to neuroinflammation and trigger of nociceptive neurons^31^. Other mechanisms that have been proposed due to COVID-19 are headache due to hypoxemia from reduced oxygenation of the CNS creating an acidic environment that then induces cerebral vasodilatation leading to headaches^32^. Several studies have also reported a propensity for ischemic events due to hypercoagulability,^33^ which may lead to headaches.

Some of the proposed reasons for varying prevalence of COVID-19-related headaches may include more risk factors that increase risk of a greater inflammatory response leading to poorer outcomes, such as sex, obesity, lower socioeconomic status or genetic predisposition^34^. Headache prevalence is known to vary in the general population by geographical regions or cultures due to differing genetic predisposition to headaches or cultural differences in pain expression^35^. For example, headache syndromes in the general population are less frequent throughout Asia compared to Latin America^36^, thus, the tendency to develop headaches in the setting of COVID-19 may differ based on these cultural, genetic or geographic differences. Next, new SARS-CoV-2 strains more prevalent in certain geographic regions may have a stronger association with particular symptoms, including headaches, compared to other variants^37^. To-date, the COVID-19 variant affecting Peru is unknown, but it may be a variant more strongly associated with headaches. There may be several reasons why worldwide prevalence of headaches associated with COVID-19 infection varies widely, and why our reported prevalence was higher than other studies in the literature.

Hyposmia and hypogeusia have been widely associated with COVID-19. Among patients with mild COVID-19 symptoms from the United States, hyposmia and hypogeusia were reported in 68% and 71%, respectively, higher than that reported in our study. Nearly two-thirds of the patients in this study reported anosmia resolution at the time of resolution of COVID-19 symptoms^38^. In one multi-center study of several European countries, a higher prevalence was reported compared with that in our study with up to 86% of patients with COVID-19 reporting anosmia or hyposmia and 88% with hypogeusia; there was a significant association between hyposmia/anosmia and hypogeusia/ageusia (p<.0001)^39^. Moreover, in up to 12%, hyposmia/anosmia or hypogeusia/ageusia was the presenting symptom^39^. In another outpatient study from Italy of patients with mild respiratory symptoms, 64% had hyposmia or anosmia^40^. Few studies from LMIC have been conducted assessing hyposmia or hypogeusia prevalence. In one study from Somalia, up to 80% of patients reported ageusia or anosmia and were the first presenting symptom in about 40% of the cohort with COVID-19 infection^41^.

In Iceland, however, anosmia prevalence was lower than that in our study affecting 11.5% of 1044 patients with COVID-19 infection^42^. Similarly, in a study from Singapore of 1065 patients with COVID-19, hyposmia or hypogeusia frequency was 12.6% with a preponderance for female sex^43^. Hyposmia or hypogeusia may be more frequently reported in outpatient settings or among those with mild-to-moderate COVID-19 due to heightened awareness of these symptoms among those who are able to continue a regular routine and diet in isolation at home, compared with hospitalized patients with more severe acute COVID-19 symptoms who may not notice or be able to report these symptoms.

However, other studies have reported prevalence of hyposmia or hypogeusia comparable to that of our study, such as in an Italian study that reported 34% of their population had either hypogeusia or hyposmia^44^. In a Canadian study, anosmia or hyposmia was more prevalent among outpatient COVID-19 patients compared with negative controls (41% vs. 5.6%), with a similar prevalence to that reported in our study^45^. In our study, hypogeusia was reported in 41% of the population and hyposmia in 40%, therefore it is possible that geographical or cultural factors, or the virus variant affecting a certain region, may play a role in the development of certain symptoms related to COVID-19.

Several mechanisms for hyposmia in the setting of COVID-19 have been proposed. Coronavirus (SARS-CoV-1), for example, has been demonstrated to spread from the olfactory epithelium along the olfactory nerve to the olfactory bulb within the CNS in a transgenic mouse model^46^. It has also been demonstrated to spread in a retrograde fashion via transsynaptic transfer of vesicles that moves the virus particles along microtubules back to the neuronal cell body^47^. The virus has a predilection for the olfactory sensory tract leading to damage of the olfactory bulbs given that angiotensin converting enzyme-2 (ACE-2) receptors, the receptors to which SARS-CoV-2 binds, are present in the olfactory epithelium^48^. MRI data suggests that obstruction of the olfactory cleft caused by transient edema in patients with severe acute SARS-CoV-2 infection during early stages of the disease may occur, but the majority of obstructions improve or resolve at 1-month follow-up^48^. Other studies have also suggested a quick recovery of olfactory disorders in the setting of COVID-19 infection with some improvement occurring in 27% of patients^49^, suggesting there is possible reversible damage to the neurosensory cells of the olfactory bulb by the virus^50^. Therefore, our findings suggest that even in those with mild respiratory symptoms, the virus may invade the olfactory epithelium in a high proportion of patients during the acute phase of illness, but there may be a potential for reversal of these symptoms as has been demonstrated in other studies.

Our study has several limitations. First, this was a cross-sectional study that captured data from participants at the time of hospital admission and did not track their clinical status over time. Thus, we do not have information on their hospital admission course, discharge or outcomes during or after hospitalization. Given the cross-sectional nature of the study, we cannot comment on whether patients presenting with mild symptoms to a public hospital in Peru have greater risk of worse outcomes if they had a neurological symptom at disease onset. Second, the study did not assess temporal patterns of neurological symptoms, thus we do not know which symptoms may have presented first and in what sequence. Third, this was a population of patients presenting with mild respiratory symptoms to a public hospital in Lima, Peru, thus we are unable to extrapolate results to other geographic regions of Peru, such as more rural regions, or to patients with moderate-to-severe symptoms of COVID-19 in Peru. Moreover, at the time of this study, rt-PCR testing was not widely available in public hospitals in Lima, Peru, thus serological antibody testing for confirmation of COVID-19 exposure was considered the most appropriate measure for diagnosis of acute COVID-19. Because we followed the diagnostic and management guidelines of public Peruvian hospitals, patients with a positive anti-SARS-CoV-2 IgM or IgG and compatible COVID-19 symptoms were considered to have acute COVID-19. Thus, there is a possibility that symptoms may have been due to co-infection with influenza or another upper respiratory viral infection, however, the large majority of patients had symptoms compatible with COVID-19 at the time of study entry rendering this possibility unlikely. In addition, our inclusion criteria only considered neurological symptoms with an onset of 14 days or less, and the mean neurological symptom duration was 8 days, thus limiting the possibility that neurological symptoms were due to another cause. Despite these limitations, we present the first study on neurological symptoms in a cohort of patients with mild-to-moderate COVID-19 in Peru.

## Conclusions

In conclusion, our study found that 83% of patients with acute mild-to-moderate COVID-19 had at least one neurological symptom during the acute infectious period, with headaches being most common (72%). We also found a high prevalence of hyposmia/anosmia (40%) and hypogeusia/ageusia (41%). In addition, factors such as non-neurological symptoms of COVID-19 (i.e. cough, dyspnea, fever) increased the risk of having at least one neurological symptom. Identifying these symptoms may be helpful particularly in Peru where molecular testing is not yet widely available in public Peruvian governmental hospitals, but has become more commonly utilized as of November 2020. Despite this, many hospitals continue to rely on antibody serological testing, thus not having an accurate count of current infection cases^23^. Identifying a person with hyposmia, hypogeusia or headache increases the likelihood of identifying a person with acute COVID-19 infection in a Peruvian population, particularly without widespread access to molecular testing throughout the country. Early screening or identification of these symptoms may be a means to identify patients who need isolate and quarantine away from household members and close contacts, even without the presence of COVID-19 respiratory symptoms. This may help mitigate spread of the infection particularly in many LMIC where COVID-19 molecular testing may not be as widely accessible.

## Data Availability

The authors can make the data set from which this study was derived to any investigator who requests it of the corresponding author.

## Acknowledgements

We would like to acknowledge the physicians of Villa Panamericana Hospital who helped in data collection, including Dr. Acxell Vásquez Moreno and Dr. Rolando Stefano Quiroz Flores.

